# Lockdown measures and relative changes in the age-specific incidence of SARS-CoV-2 in Spain

**DOI:** 10.1101/2020.06.30.20143560

**Authors:** PM De Salazar, D Gómez-Barroso, D Pampaka, JM Gil, B Peñalver, C Fernández-Escobar, M Lipsitch, A Larrauri, E Goldstein, MA Hernán

**Author notes:** Co-senior authors (E Goldstein, MA Hernán).

## Abstract

**Background:** The first months of the SARS-CoV-2 epidemic in Spain resulted in high incidence and mortality. A national sero-epidemiological survey suggests higher cumulative incidence of infection in older individuals than in younger individuals. However, little is known about the epidemic dynamics in different age groups, including the relative effect of the lockdown measures introduced on March 15, and strengthened on March 30 to April 14, 2020 when only essential workers continued to work.

**Methods:** We used data from the National Epidemiological Surveillance Network (RENAVE in Spanish) on the daily number of reported COVID-19 cases (by date of symptom onset) in eleven 5-year age groups: 15-19y through 65-69y. For each age group g, we computed the proportion E(g) of individuals in age group g among all reported cases aged 15-69y during the pre-lockdown period (March 1-10, 2020) and the corresponding proportion L(g) during two lockdown periods (March 25-April 3 and April 8-17, 2020). For each lockdown period, we computed the proportion ratios PR(g)= L(g)/E(g). For each pair of age groups g1,g2, PR(g1)>PR(g2) implies a relative increase in the incidence of detected SARS-CoV-2 infection in the age group g1 compared with g2 for the later vs. early period.

**Results:** For the first lockdown period, the highest PR values were in age groups 50-54y (PR=1.21; 95% CI: 1.12,1.30) and 55-59y (PR=1.19; 1.11,1.27). For the second lockdown period, the highest PR values were in age groups 15-19y (PR=1.26; 0.95,1.68) and 50-54y (PR=1.20; 1.09,1.31).

**Conclusions:** Our results suggest that different outbreak control measures led to different changes in the relative incidence by age group. During the first lockdown period, when non-essential work was allowed, individuals aged 40-64y, particularly those aged 50-59y presented with higher COVID-19 relative incidence compared to pre-lockdown period, while younger adults/older adolescents (together with persons aged 50-59y) had increased relative incidence during the later, strengthened lockdown. The role of different age groups during the epidemic should be considered when implementing future mitigation efforts.

## Introduction

The ongoing SARS-CoV-2 epidemic led to the implementation of mitigation strategies worldwide. To understand SARS-CoV-2 dynamics under various mitigation strategies, it is important to study the role of age because variations in transmission by age suggest differential impact of various measures such as physical distancing and workforce-related policies, which in turn has implications for epidemic control.

The rates of SARS-CoV-2 infection seem to vary by age. Serological studies in England [1], Switzerland [2] and Germany [3] found highest infection rates in younger adults and older adolescents. A serological study in New York State found comparable rates of infection in different age groups of adults in the greater New York City area, with lower rates of infection in persons aged over 55y compared with younger adults outside the greater New York City area [4]. A serological study in Spain found higher rates of infection in older individuals compared with younger ones [5].

Control measures may have differential effectiveness in different age groups [6], and the groups for which the measures are more effective may vary across populations. Social distancing seems to have been less effective among younger adults and older adolescents in Germany [7] and among persons aged 50-59 years in the Netherlands [8].

In Spain, a national lockdown was instituted on March 15 and was further strengthened to include work restrictions to non-essential workers between March 30 - April 14 [8]. This intervention had a pronounced effect on transmission that led to decreasing case counts in all regions of the country. However, little information is available on the relative incidence of different age groups during the different stages of the lockdown. For example, age groups that were overrepresented among those who continued non-remote work between March 15-29 could have had their relative incidence increase during that period.

Here, we estimate changes in detected incidence of SARS-CoV-2 infection by age group after the implementation of the different lockdown measures. We applied the methodology developed earlier [7,10,11] to assess the changes in the incidence of different age groups of individuals between the age of 15-69y during the epidemic associated with the implementation of the control measures on March 15 and March 30 in Spain.

## Methods

### Data sources

Information on daily COVID-19 cases by age group was obtained from the National Epidemiological Surveillance Network (RENAVE) through the Web platform SiViEs (System for Surveillance in Spain) through April 28, 2020 (see Figure 1, Results). We retrieved data on reported PCR-confirmed cases with available information on the date of symptom onset. We excluded healthcare workers due to significant non-community transmission in that population group. The data was retrieved on May 20, which greatly reduces right-censoring.

**Figure 1:**
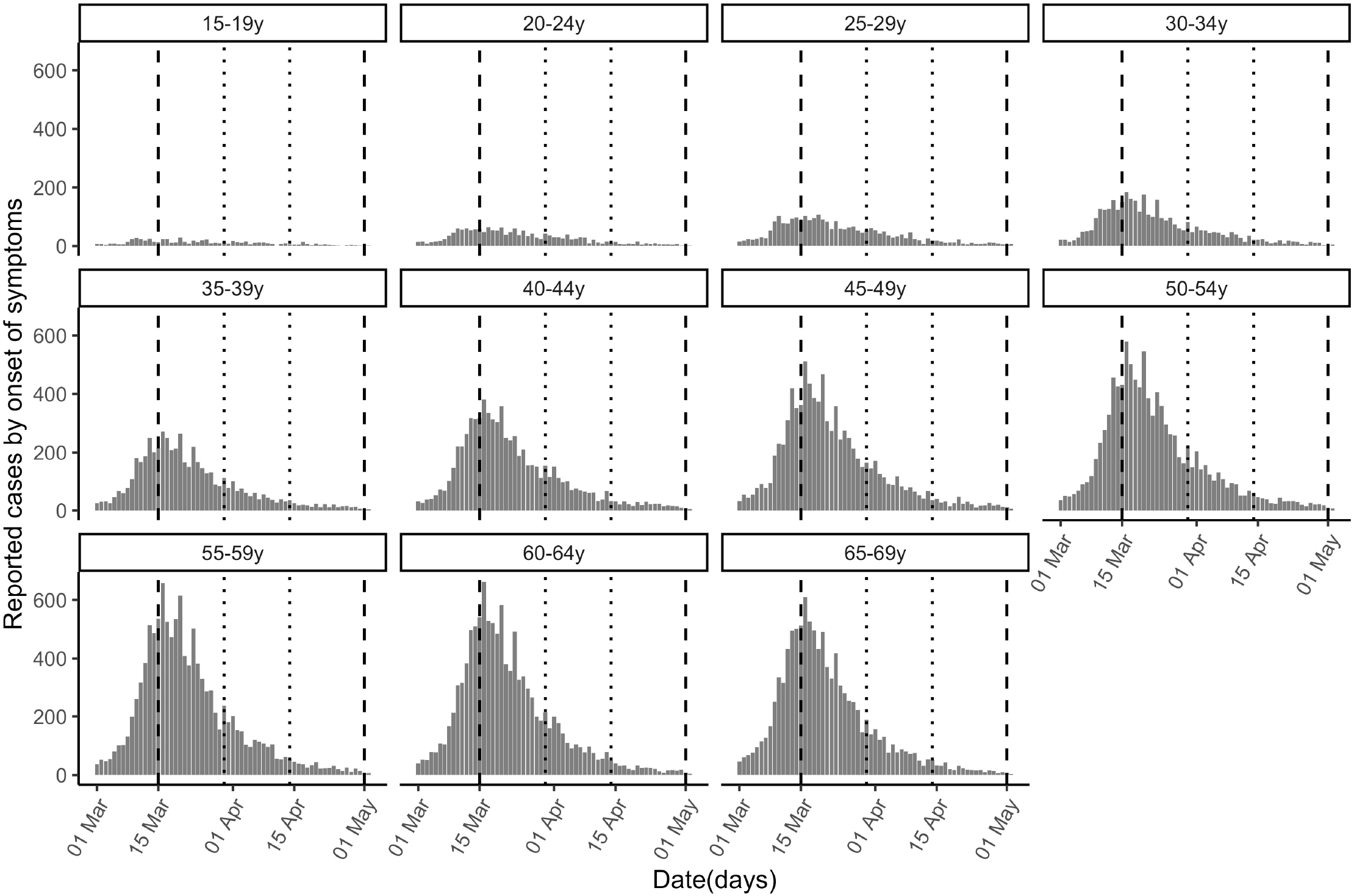
Cases of COVID-19 (reported by the day of symptom onset) by age group between March 1 and April 30, 2020 in Spain. Vertical dashed lines demarcate the first lockdown period (March 15 to April 30) and dotted lines the second (strengthened) lockdown period (March 30-April 14).

### Relative change in SARS-CoV-2 infection by age-group

Daily counts (by date of symptom onset) of reported COVID-19 cases in different 5-year age groups are plotted in Figure 1. For this analysis, we included individuals aged 15 through 69 years. We excluded children under 15 years because of potential temporal changes in diagnosis/ascertainment of cases, and individuals 70 years and older because significant non-community transmission of infection in long-term care facilities (LTCFs) likely affected their relative share in cases [13,13]. In sensitivity analyses we restricted the calculations to a) two regional clusters to explore geographic heterogeneity, and b) hospitalized cases to evaluate the impact of potential changes in ascertainment, under the assumption that the detection of severe cases requiring hospitalization is relatively insensitive to changes in diagnostic criteria.

We selected three periods (by symptom onset date of cases): March 1-10 (five days before the national lockdown, as some social distancing measure were already being phased in starting March 10), March 25-April 3 (starting ten days after the start of the lockdown to allow for time from infection to symptom onset, as well as the time for the transmission dynamics under the new set of mitigation efforts to take shape), and April 8-17 (starting 10 days after the strengthened lockdown). During the initial lockdown, but not during the strengthened lockdown, non-essential workers were allowed to commute (and work) when remote working was not possible [9].

Using the methodology in [7], we computed the age-specific proportion ratios for each of the lockdown periods (March 25-April 3 or April 8-17) relative to the pre-lockdown period (March 1-10). For each age group *g*, let *E(g)* be the number of detected COVID-19 cases in age group *g* during the earlier period (March 1-10), and *L(g)* be the corresponding number during the later period (either March 25-April 3 or April 8-17). The proportion ratio (PR) statistic is

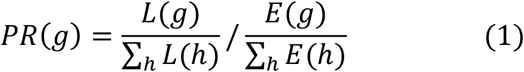

Specification of the confidence bounds for the PR statistic [14], as well as the comparison of proportion ratios in different age groups are described in section S3 of the Supplementary Material.

## Results

Figure 1 plots the epidemic curves of daily (by the day of symptom onset) COVID-19 cases for eleven 5-year age groups: (15-19y through 65-69y) between March 1^st^ and April 30^th^, 2020. Table S4 in the Supplementary Material summarizes the number of cases reported by age group for each period used in the analysis. The counts of confirmed cases are much higher in older individuals than in younger ones; however, those differences do not necessarily reflect differences in the rates of infection (as suggested by the serological estimates [5]) as infections are more severe in older individuals, and the likelihood of reporting of infection is higher for older individuals than for younger ones.

### After first lockdown period: March 25-April 3, 2020

Figure 2 plots the estimates of the proportion ratio (PR) for the period of March 25–April 3 vs. March 1^st^–March 10. Among the age groups considered, the highest estimates of PR belong to persons aged 50-54y (PR=1.21; 95% CI 1.12,1.30) and 55-59y (PR=1.19; 1.11,1.27), with PR estimates for persons aged 15-44y and 60-69y being significantly lower (Supplementary Material).

**Figure 2:**
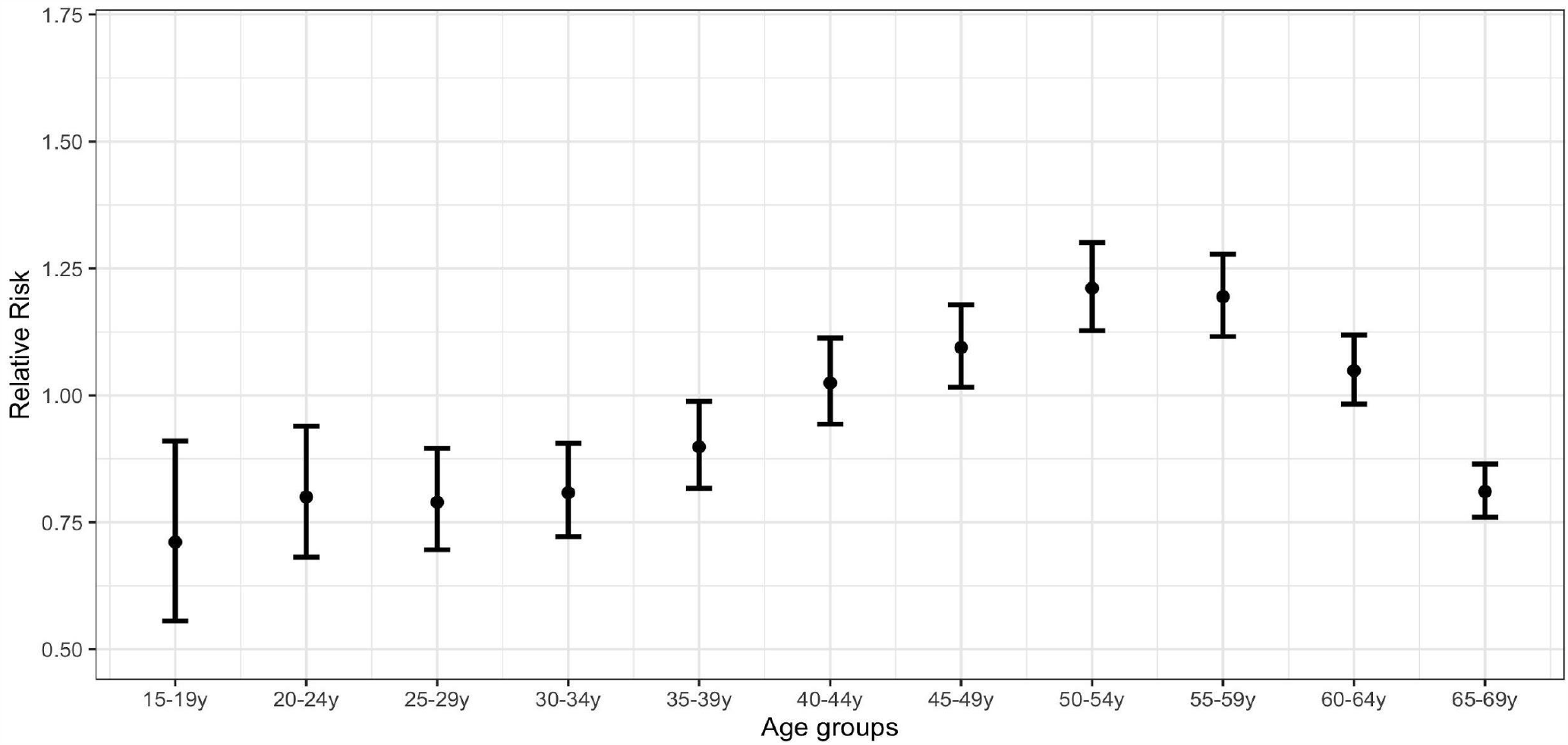
Proportion ratio (PR) estimates of confirmed COVID-19 cases by age group in Spain for the period March 25–April 3 vs. March 1-10.

### After second (strengthened) lockdown period: April 8-17, 2020

Figure 3 plots the estimates of PR for the period of April 8-17 vs. March 1-10, 2020. Among the age groups considered, the highest point estimate of PR belongs to individuals aged 15-19y (PR=1.26; 0.95,1.68), followed by persons aged 50-54y (PR=1.20; 1.09,1.31), 55-59y (PR=1.16; 1.06,1.27), and 30-34y (PR=1.08; 0.94,1.25).

**Figure 3:**
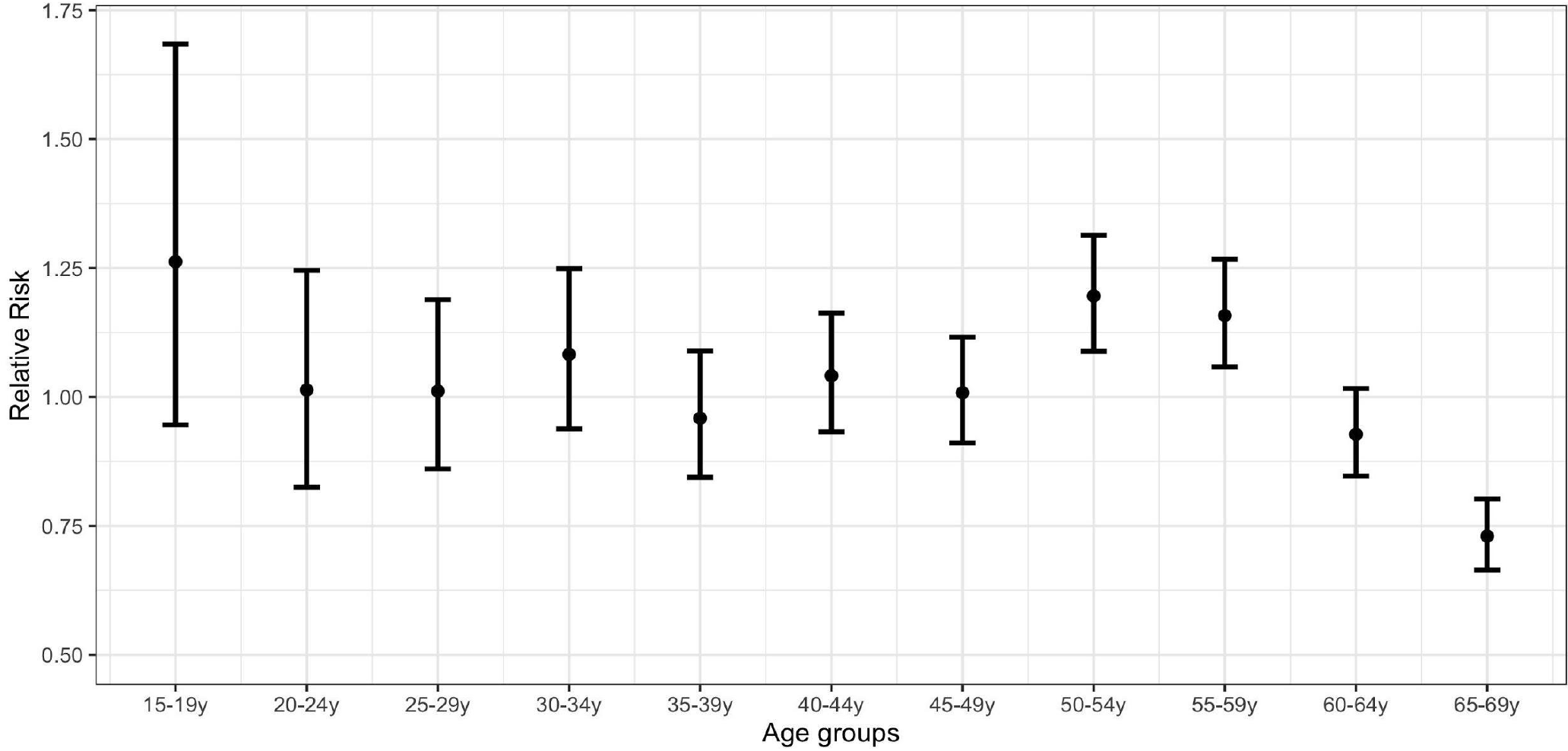
Proportion ratio (PR) estimates of confirmed COVID-19 cases by age group in Spain for the period April 8-17 vs. March 1-10.

A comparison of Figures 2 and 3 suggests an increase in the proportion of COVID-19 cases for the second compared with the first lockdown period in younger age groups (up to 34 years) relative to the middle ones (35-64y). Those increases for different pairs of age groups are shown in Table S3 in the Supplementary Material.

Sensitivity analyses using regional clusters and hospitalized cases yielded consistent estimates (Supplementary Material, Sections S1-S2).

## Discussion

We applied the methodology in [7,10,11] to show that the relative incidence of detected SARS-CoV-2 infections in different age groups changed during the lockdown periods in Spain. Individuals aged 40-64y, particularly those aged 50-59y, had an elevated relative incidence during the first lockdown period, when non-essential work was allowed, compared to pre-lockdown period. The relative incidence of younger adults/older adolescents, as well as persons aged 50-59y, was increased during a strengthened lockdown compared to pre-lockdown period.

These differences by age group might be explained by a number of factors. First, the elevated relative incidence in middle-aged adults during the first lockdown period, when non-essential workers were allowed to work, is consistent with the higher employment rates in Spain in middle-aged adults compared with younger adults [15]. Second, adherence to social distancing may vary with age: relative increases for younger adults/older adolescents during the second lockdown period might reflect lower adherence to lockdown measures, whereas perception for risk of severe disease could have led to stronger individual adherence for persons aged 60-69y. Finally, changing social responsibilities, household transmission, and the high prevalence of multigenerational families may also have contributed to the patterns of incidence observed in this paper. Further work is needed to understand those issues to better inform future mitigation efforts.

Our results are aligned with findings in several other European countries. Persons aged 50-59y were least impacted in terms of mixing/number of contacts with people following the introduction of social distancing measures in the Netherlands [8], which is in agreement with the elevated relative incidence in persons (excluding healthcare workers) aged 50-59y throughout the lockdown period in Spain. Our findings for the second lockdown period in Spain echo those from several other countries: a higher relative incidence of infection in younger persons was found in England [1], Switzerland [2], and Germany (Figure 6A in [3]), and the highest proportion ratio (PR) estimates for the post lockdown period in Germany belong to younger persons [7].

Our findings could be affected by age-differential changes in case ascertainment, over time or across regions. However, there is no evidence of fundamental diagnostic changes during the lockdown period in Spain, where the focus was on testing the more severe cases. Moreover, analyses restricted to hospitalized cases, which are less likely affected by changes in ascertainment, and region-specific analyses yielded estimates that were consistent with those of the main analysis.

In summary, our paper provides evidence for an elevated relative incidence of individuals aged 40-64y during the 1^st^ lockdown period, when non-essential work was allowed, and for an elevated relative incidence of younger adults/older adolescents when only essential workers continued working. These results suggest that age structure is an important factor in the effect of lockdown interventions.

## Data Availability

NA

## Acknowledgements

The authors of this paper would like to thank all the contributors to the surveillance and investigation of COVID-19 epidemic in Spain. This includes colleagues at local and regional Public Health Services, as well as the Centre of Health Alerts and Emergencies at the Ministry of Health, CNE and the National Microbiology Centre of the Carlos III Institute of Health.

We are particularly grateful to all the healthcare workers, for their resilience in providing treatment and care to patients of COVID-19, conducting tests and reporting the best possible data in this emergency.

## Supplementary Material

### Section S1: Proportion ratios by geographical cluster

A hierarchical cluster analysis (Figure S1) classified Spanish regions (“comunidades autónomas”) in two clusters: a central cluster (around Madrid), which has higher serologically confirmed incidence rates [2], and a peripheral cluster with lower incidence (Figure S2).

**Figure S1:**
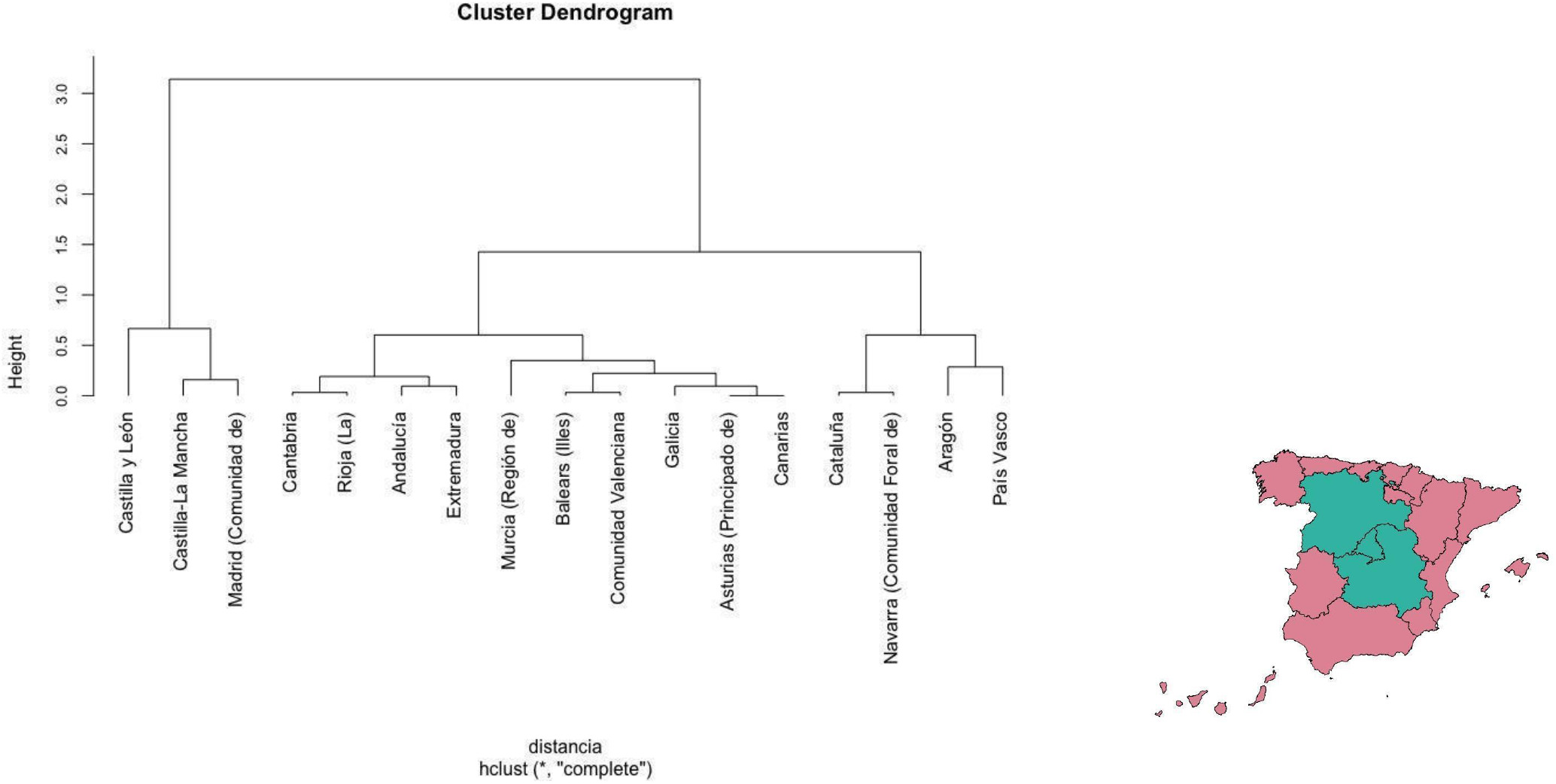
Splitting of Spain into a Central cluster and Peripheral cluster.

**Figure S2:**
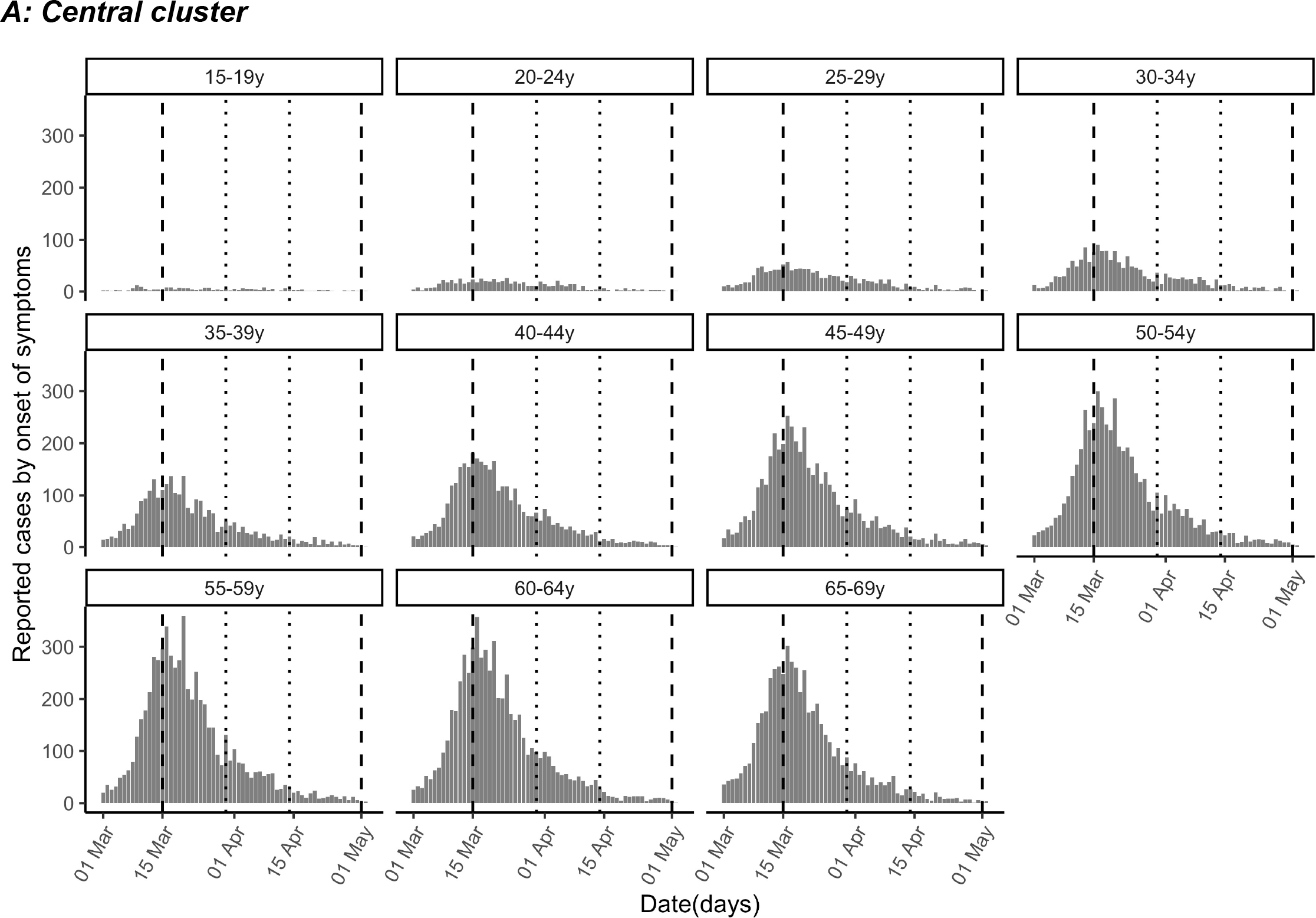
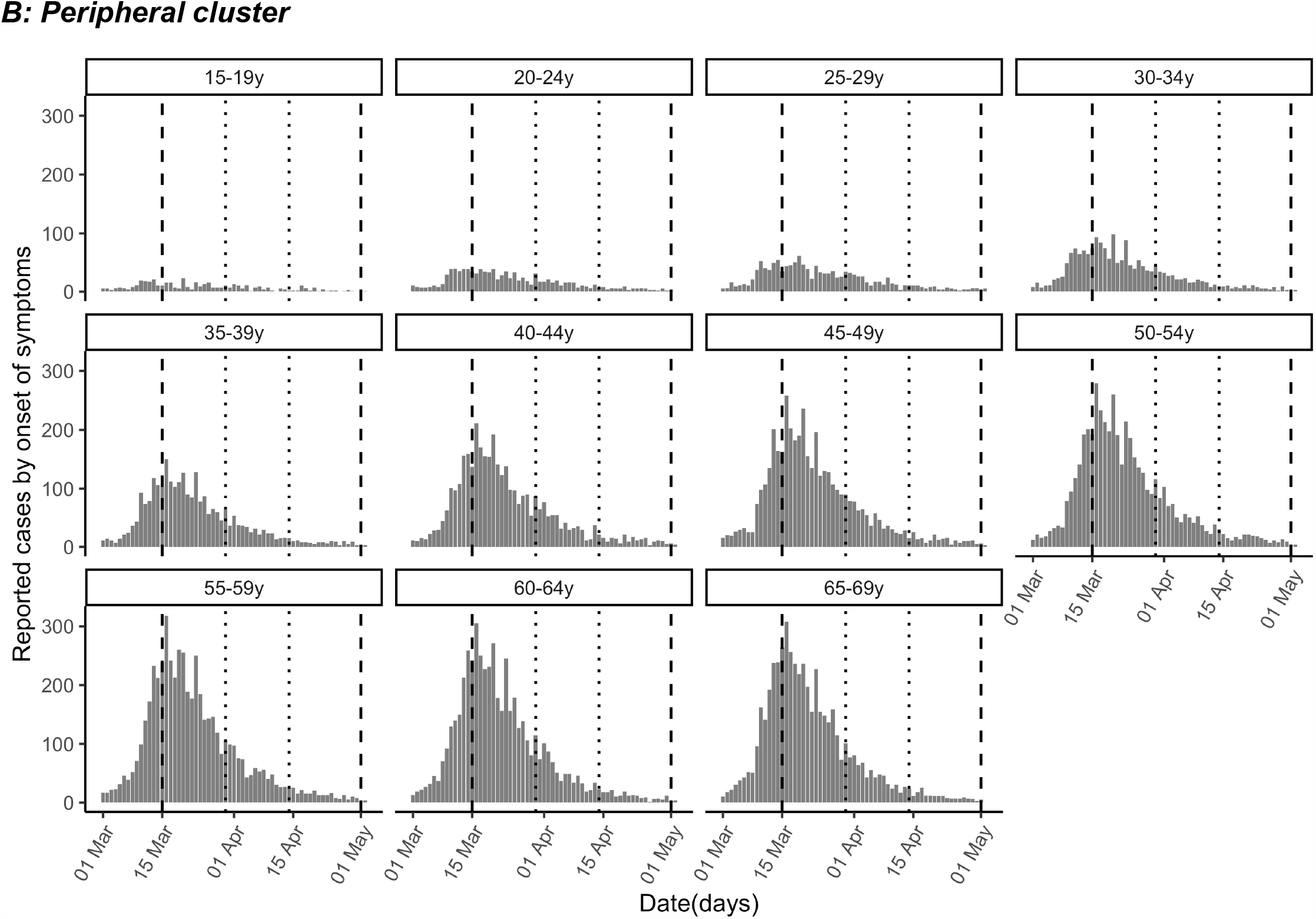
Cases of COVID-19 (reported by the day of symptom onset) by age group between March 1 and April 30, 2020 in each the Central cluster (A) and Peripheral cluster (B) in Spain. Vertical dashed lines demarcate the first lockdown period (March 15 to April 30) and dotted lines the second (strengthened) lockdown period (March 30-April 14).

**Figure S3:**
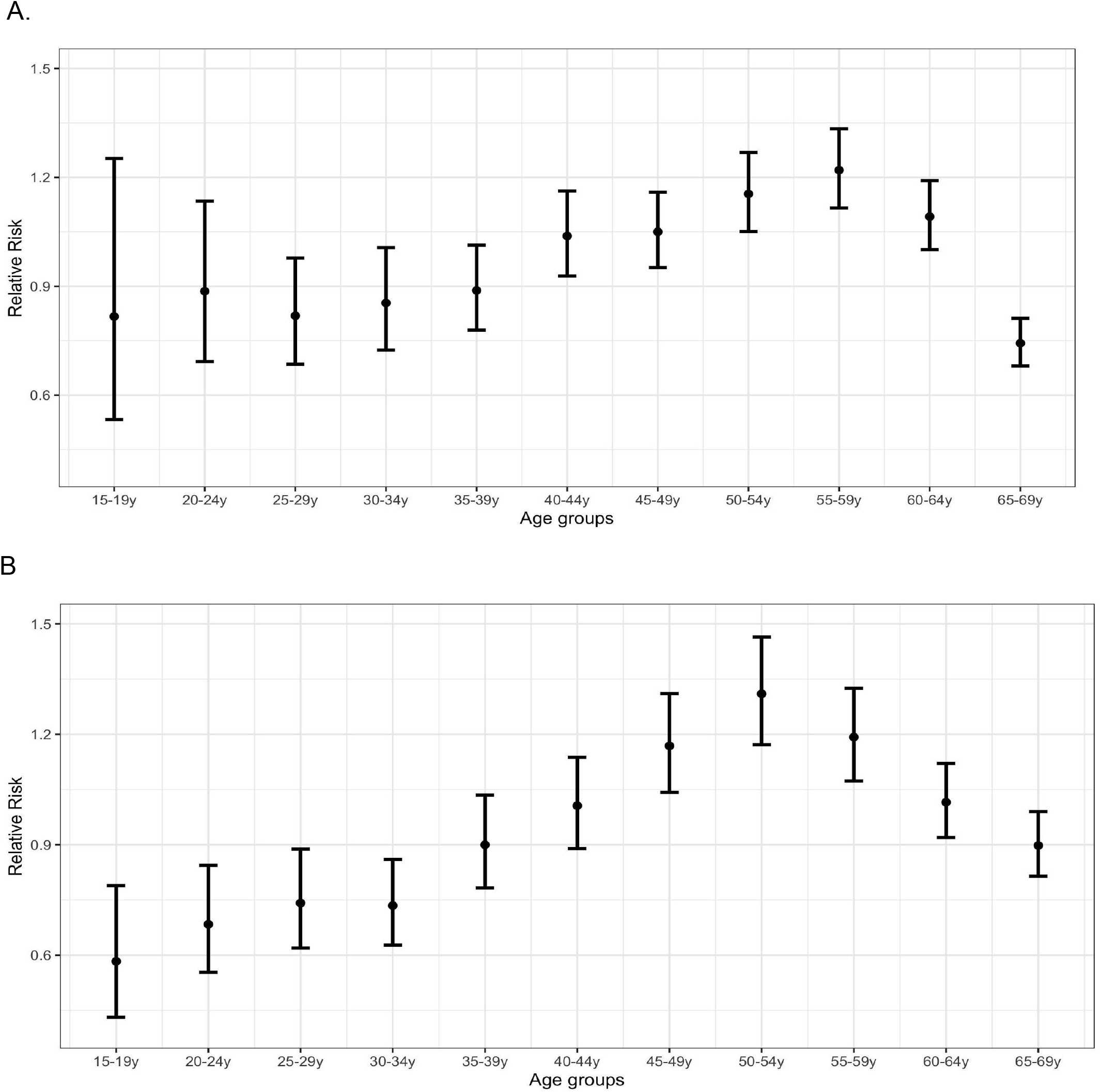
Proportion ratio (PR) estimates of confirmed COVID-19 cases by age group in Spain for the period March 25–April 3 vs. March 1-10 for the Central cluster (A) and the Peripheral cluster (B).

**Figure S4:**
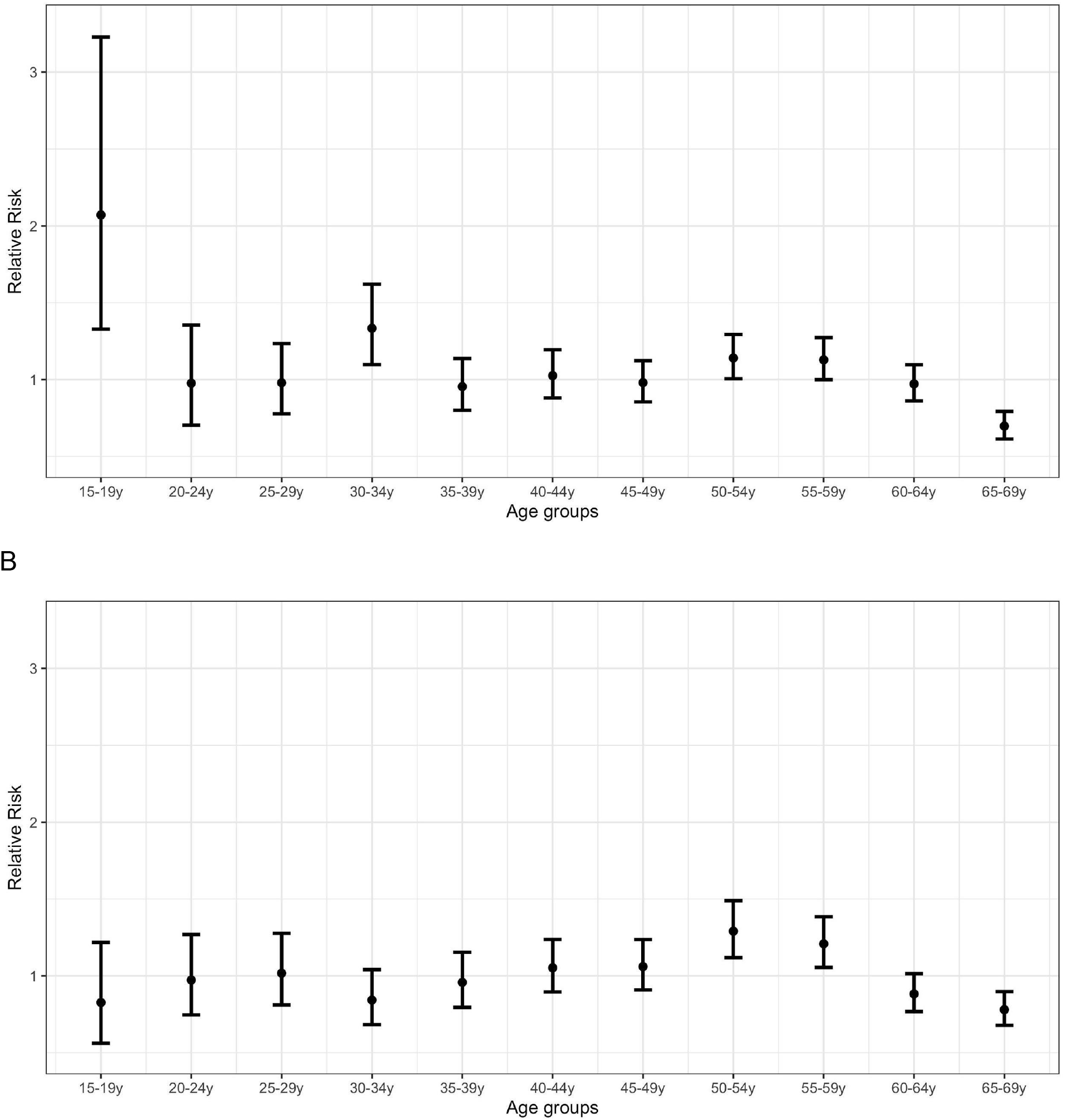
Proportion ratio (PR) estimates of confirmed COVID-19 cases by age group in Spain for the period April 8-17 vs. March 1-10 for the Central cluster (A) and the Peripheral cluster (B).

### Section S2: Proportion ratio by age-group for hospitalized cases

Figure S5A plots the estimates of the proportion ratio (PR) for the period of March 25 –April 3 vs. March 1 –March 10 (eq. 1) and Figure S5B plots PR for the period April 8-17 vs March 1-March 10 for individuals requiring hospitalization (n=72283, obtained from RENAVE /SiViEs for those with available information, see Methods). While the confidence bounds in Figures S5A, B are wide due to smaller sample size, especially within younger age groups, the point estimates shown in Figure S5A are in alignment to those shown in Figure 2 with the highest PR estimates belonging to persons aged 50-59y. Similarly, the estimates showed in Figure S5B are consistent with those in Figure 3, with a relative increase in old adolescents/younger adults (15-34y) in addition to those 50-59y.

**Figure S5:**
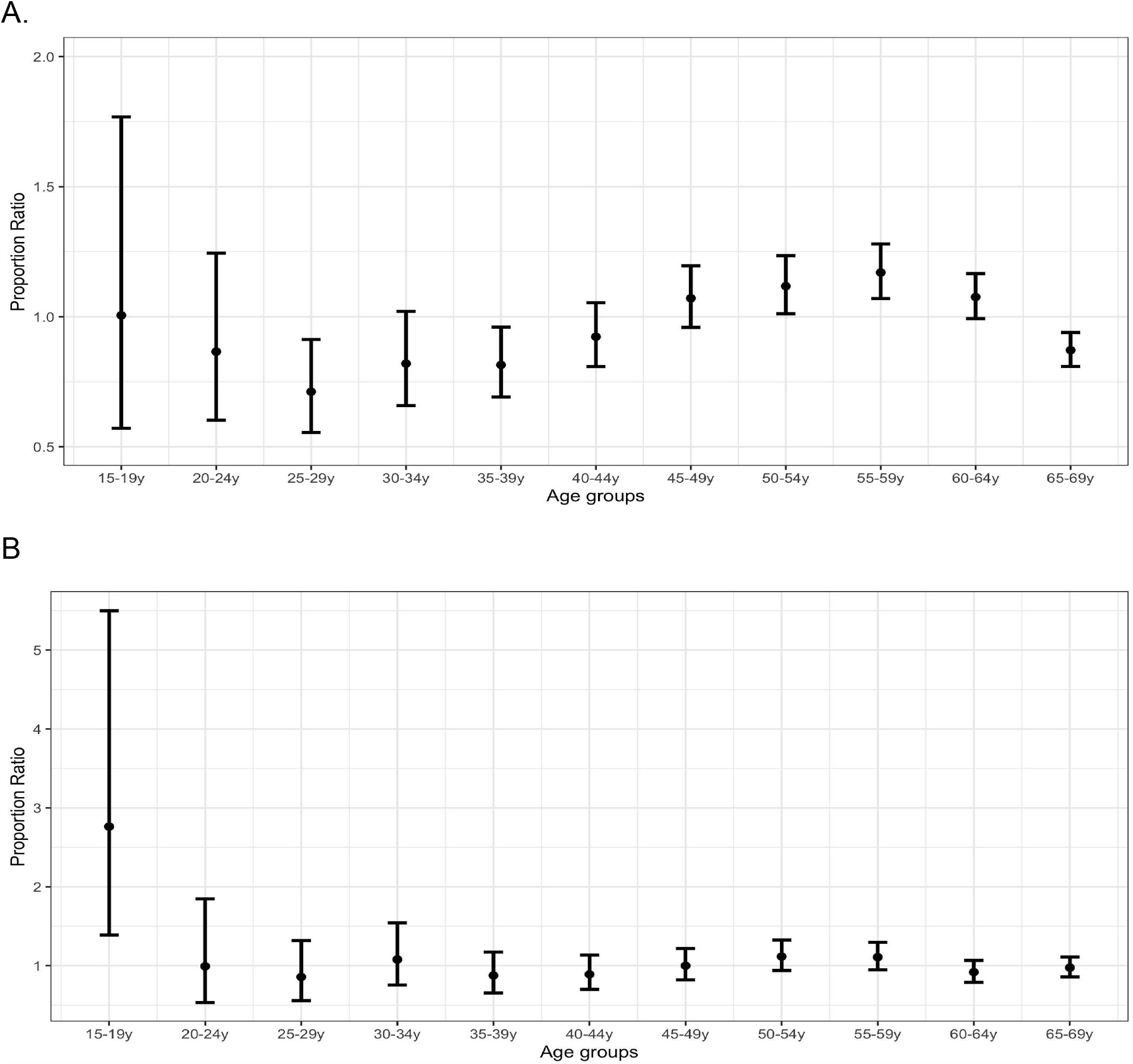
Estimates of the proportion ratio (PR) for hospitalized cases the period of March 25 –April 3 vs. March 1st –March 10 (A) and April 8 –April 17 vs. March 1st –March 10

### Section S3: Confidence bounds and pairwise comparison for proportion ratios

For the proportion ratio statistic introduced in the main text (eq. 1), the logarithm ln(*PR*(*g*)) of the PR(*g)* is approximately normally distributed [14] with the standard error:

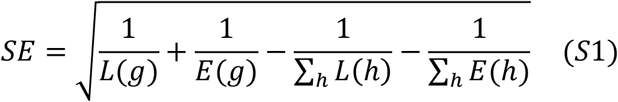

For each pair of age groups *g*1 and *g*2, the proportion ratios *PR*(*g*1) and *PR*(*g*2) are compared using the odds ratio (OR)

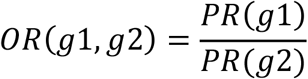

It follows from eq. 1 in the main body of the text that *OR*(*g*1,*g*2) equals

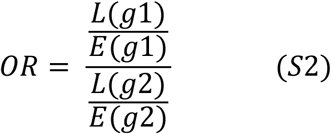

which is the OR for a COVID-19 case to be in age group *g*1 vs *g*2 for the later vs early period. Estimates for pairwise OR were performed using Fisher’s exact test. Table S1 gives the estimates of the odds ratios (ORs) for different pairs of age groups (15-29y through 65-69y) for a COVID-19 case to be detected for the period of March 25 –April 3 vs. March 1st –March 10 (eq. 3). Table S1 suggests that for persons aged 50-59y, the corresponding odds ratio relative to any age group either between 15-39y, or above 60y is significantly above 1. For persons aged 45-49y, the corresponding odds ratio relative to any age group either between 15-39y, or 65-69y is significantly above 1.

**Table S1.** Odds ratios (ORs) for different pairs of age groups (15-19y through 65-69y) for a COVID-19 case to be detected for the period of March 25 –April 3 vs. March 1st –March 10 (eq. 3).

| Age group (years ) | 20-24 | 25-29 | 30-34 | 35-39 | 40-44 | 45-49 | 50-54 | 55-59 | 60-64 | 65-69 |
| --- | --- | --- | --- | --- | --- | --- | --- | --- | --- | --- |
| 15-19 | 0.89<br>(0.65-1.21) | 0.9<br>(0.6-1.20) | 0.88<br>(0.61-1.17) | 0.79<br>(0.57-1.04) | **0.69**<br>**(0.53-0.91)** | **0.65**<br>**(0.5-0.85)** | **0.59**<br>**(0.45-0.77)** | **0.60**<br>**(0.46-0.78)** | **0.68**<br>**(0.52-0.89)** | 0.88<br>(0.67-1.15) |
| 20-24 |  | 1.01<br>(0.82-1.25) | 0.99<br>(0.81-1.22) | 0.89<br>(0.73-1.08) | **0.78**<br>**(0.65-0.94)** | **0.73**<br>**(0.61-0.88)** | **0.66**<br>**(0.55-0.79)** | **0.67**<br>**(0.56-0.80)** | **0.76**<br>**(0.64-0.92)** | 0.99<br>(0.82-1.18) |
| 25-29 |  |  | 0.98<br>(0.82-1.17) | 0.88<br>(0.74-1.04) | **0.77**<br>**(0.66-0.90)** | **0.72**<br>**(0.62-0.84)** | **0.65**<br>**(0.56-0.76)** | **0.66**<br>**(0.57-0.77)** | **0.75**<br>**(0.65-0.87)** | 0.97<br>(0.84-1.13) |
| 30-34 |  |  |  | 0.90<br>(0.77-1.05) | **0.79**<br>**(0.68-0.92)** | **0.74**<br>**(0.64-0.85)** | **0.67**<br>**(0.58-0.77)** | **0.68**<br>**(0.59-0.78)** | **0.77**<br>**(0.67-0.89)** | 1.0<br>(0.87-1.15) |
| 35-39 |  |  |  |  | 0.88<br>(0.77-1) | **0.82**<br>**(0.72-0.93)** | **0.74**<br>**(0.65-0.84)** | **0.75**<br>**(0.66-0.85)** | **0.86**<br>**(0.76-0.97)** | 1.11<br>(0.98-1.25) |
| 40-44 |  |  |  |  |  | 0.94<br>(0.83-1.05) | **0.85**<br>**(0.75-0.95)** | **0.86**<br>**(0.76-0.96)** | 0.98<br>(0.87-1.09) | **1.26**<br>**(1.13-1.42)** |
| 45-49 |  |  |  |  |  |  | 0.90<br>(0.81-1.01) | 0.92<br>(0.82-1.02) | 1.04<br>(0.94-1.16) | **1.35**<br>**(1.21-1.50)** |
| 50-54 |  |  |  |  |  |  |  | 1.03<br>(0.9-1.15) | **1.15**<br>**(1.04-1.28)** | **1.49**<br>**(1.34-1.66)** |
| 55-59 |  |  |  |  |  |  |  |  | **1.14**<br>**(1.03-1.26)** | **1.47**<br>**(1.33-1.63)** |
| 60-64 |  |  |  |  |  |  |  |  |  | **1.29**<br>**(1.17-1.43)** |

Table S2 shows the estimates of the odds ratios (ORs) for different pairs of age groups (15-19y through 65-69y) for a COVID-19 case to be detected for the period of April 8-17 vs. March 1st –March 10 (eq. 3). Table S2 suggests that for persons aged 40-59y, the corresponding odds ratio relative to persons aged 60-69y is significantly above 1.

**Table S2.** Odds ratios (ORs) for different pairs of age groups (15-19y through 65-69y) for a COVID-19 case to be detected for the period of April 8-17 vs. March 1st –March 10 (eq. 3).

| Age group (years) | 20-24 | 25-29 | 30-34 | 35-39 | 40-44 | 45-49 | 50-54 | 55-59 | 60-64 | 65-69 |
| --- | --- | --- | --- | --- | --- | --- | --- | --- | --- | --- |
| 15-19 | 1.27<br>(0.9-1.79) | 1.33<br>(0.97-1.83) | 1.27<br>(0.93-1.74) | **1.42**<br>**(1.05-1.92)** | 1.32<br>(0.98-1.77) | 1.34<br>(1-1.80) | 1.15<br>(0.86-1.55) | 1.22<br>(0.91-1.63) | **1.55**<br>**(1.15-2.07)** | **1.95**<br>**(1.45-2.61)** |
| 20-24 |  | 1.05<br>(0.81-1.35) | 1.<br>(0.79-1.28) | 1.12<br>(0.88-1.41) | 1.04<br>(0.83-1.30) | 1.06<br>(0.85-1.32) | 0.91<br>(0.73-1.13) | 0.96<br>(0.77-1.19) | 1.22<br>(0.98-1.51) | **1.53**<br>**(1.23-1.91)** |
| 25-29 |  |  | 0.96<br>(0.78-1.18) | 1.07<br>(0.87-1.30) | 0.99<br>(0.82-1.20) | 1.01<br>(0.84-1.22) | 0.87<br>(0.72-1.04) | 0.91<br>(0.76-1.10) | 1.16<br>(0.97-1.40) | **1.47**<br>**(1.22-1.76)** |
| 30-34 |  |  |  | 1.11<br>(0.92-1.34) | 1.03<br>(0.86-1.23) | 1.05<br>(0.88-1.25) | 0.9<br>(0.76-1.07) | 0.95<br>(0.81-1.13) | **1.21**<br>**(1.02-1.44)** | **1.53**<br>**(1.28-1.82)** |
| 35-39 |  |  |  |  | 0.93<br>(0.78-1.10) | 0.95<br>(0.8-1.11) | **0.81**<br>**(0.69-0.95)** | **0.86**<br>**(0.73-1.00)** | 1.09<br>(0.93-1.28) | **1.37**<br>**(1.17-1.61)** |
| 40-44 |  |  |  |  |  | 1.02<br>(0.88-1.19) | **0.88**<br>**(0.76-1.01)** | 0.92<br>(0.8-1.07) | **1.18**<br>**(1.02-1.36)** | **1.48**<br>**(1.28-1.72)** |
| 45-49 |  |  |  |  |  |  | **0.86**<br>**(0.75-0.99)** | 0.91<br>(0.79-1.04) | **1.15**<br>**(1.00-1.32)** | **1.45**<br>**(1.26-1.67)** |
| 50-54 |  |  |  |  |  |  |  | 1.05<br>(0.92-1.21) | **1.34**<br>**(1.17-1.53)** | **1.69**<br>**(1.48-1.94)** |
| 55-59 |  |  |  |  |  |  |  |  | **1.27**<br>**(1.11-1.45)** | **1.60**<br>**(1.40-1.83)** |
| 60-64 |  |  |  |  |  |  |  |  |  | **1.26**<br>**(1.10-1.44)** |

Table S3 shows the estimates of the odds ratios (ORs) for different pairs of age groups (15-19y through 65-69y) for a COVID-19 case to be detected for the period of April 8-17 vs. March 25-April 3 (eq. 3). Table S3 shows an increase in the incidence of detected SARS-CoV-2 infection for younger persons (aged under 34y) compared to older persons (aged over 40y) for a number of pairs of age groups.

**Table S3.** Odds ratios (ORs) for different pairs of age groups (15-19y through 65-69y) for a COVID-19 case to be detected for the period of April 8 –April 17 vs. March 25 –April 3 (eq. 3).

| Age group (years ) | 20-24 | 25-29 | 30-34 | 35-39 | 40-44 | 45-49 | 50-54 | 55-59 | 60-64 | 65-69 |
| --- | --- | --- | --- | --- | --- | --- | --- | --- | --- | --- |
| 15-19 | 1.4<br>(0.98-2) | 1.39<br>(0.99-1.93) | 1.33<br>(0.96-1.83) | 1.66<br>(1.21-2.28) | 1.75<br>(1.28-2.38) | 1.93<br>(1.41-2.61) | 1.8<br>(1.32-2.43) | 1.83<br>(1.35-2.47) | 2.01<br>(1.47-2.72) | 1.99<br>(1.45-2.67) |
| 20-24 |  | 0.99<br>(0.76-1.28) | 0.95<br>(0.74-1.21) | 1.25<br>(0.93-1.51) | 1.25<br>(0.99-1.57) | 1.37<br>(1.09-1.72) | 1.28<br>(1.02-1.60) | 1.31<br>(1.04-1.63) | 1.43<br>(1.14-1.79) | 1.41<br>(1.12-1.76) |
| 25-29 |  |  | 0.96<br>(0.77-1.19) | 1.20<br>(0.98-1.47) | 1.26<br>(1.04-1.53) | 1.39<br>(1.15-1.68) | 1.30<br>(1.08-1.56) | 1.32<br>(1.1-1.59) | 1.45<br>(1.2-1.74) | 1.42<br>(1.18-1.72) |
| 30-34 |  |  |  | 1.25<br>(1.04-1.52) | 1.32<br>(1.1-1.58) | 1.45<br>(1.22-1.73) | 1.36<br>(1.14-1.61) | 1.38<br>(1.17-1.63) | 1.51<br>(1.28-1.79) | 1.49<br>(1.25-1.77) |
| 35-39 |  |  |  |  | 1.05<br>(0.89-1.24) | 1.16<br>(0.98-1.36) | 1.08<br>(0.92-1.26) | 1.10<br>(0.94-1.28) | 1.21<br>(1.03-1.41) | 1.18<br>(1.01-1.39) |
| 40-44 |  |  |  |  |  | 1.10<br>(0.95-1.28) | 1.03<br>(0.89-1.19) | 1.05<br>(0.91-1.21) | 1.15<br>(1-1.33) | 1.13<br>(0.97-1.31) |
| 45-49 |  |  |  |  |  |  | 0.93<br>(0.81-1.07) | 0.95<br>(0.83-1.09) | 1.04<br>(0.91-1.19) | 1.02<br>(0.89-1.18) |
| 50-54 |  |  |  |  |  |  |  | 1.02<br>(0.9-1.16) | 1.12<br>(0.98-1.27) | 1.10<br>(0.96-1.25) |
| 55-59 |  |  |  |  |  |  |  |  | 1.10<br>(0.96-1.25) | 1.08<br>(0.94-1.23) |
| 60-64 |  |  |  |  |  |  |  |  |  | 0.98<br>(0.86-1.12) |

### Section S4: Number of reported cases in different age groups during different time periods

Table S4 shows the number of reported COVID-19 cases with available information on the date of symptom onset in different age groups for different time periods. Those case counts increased a great deal with age, likely reflecting higher case ascertainment for older individuals.

**Table S4:** Number of reported COVID-19 cases with available information on the date of symptom onset in different age groups for different time periods.

| Period | 15-19y | 20-24y | 25-29y | 30-34y | 35-39y | 40-44y | 45-49y | 50-54y | 55-59y | 60-64y | 65-69y |
| --- | --- | --- | --- | --- | --- | --- | --- | --- | --- | --- | --- |
| March 1-10 | 113 | 250 | 399 | 482 | 646 | 795 | 942 | 970 | 1065 | 1200 | 1342 |
| March 25 - April 3 | 139 | 346 | 545 | 674 | 1004 | 1409 | 1783 | 2032 | 2200 | 2177 | 1882 |
| April 8 -17 | 76 | 135 | 215 | 278 | 330 | 441 | 506 | 618 | 657 | 593 | 522 |

